# Reproducible variability: assessing human vs. AI discordance in attempting to reproduce the same observational study

**DOI:** 10.64898/2026.11.25.26369008

**Authors:** Joshua Walonoski, Chloe W. Eng, Armaan Singh Johal, Alex Bennett, Max Fleming, Yorick Chern

## Abstract

**Objective:** Previously, nine expert teams given a published study and identical code sets produced nine substantially different cohorts, prompting the conclusion that analytic code cannot be reliably generated from natural-language description. We tested whether an artificial intelligence (AI) agent, given that same description, reproduces a cohort definition more faithfully and consistently than those teams.

**Materials and Methods:** Against a claims database of 35.9 million patients, we executed 27 independent reproductions of that published glucagon-like peptide-1 receptor agonist cohort definition: the nine published human definitions re-implemented deterministically (Arm A); nine independent AI agent runs given the source paper as published (Arm B); and nine given a rewrite that removed its internal contradictions (Arm C). All arms received identical frozen code sets. Agreement with a reference cohort was quantified by the Jaccard index.

**Results:** Agents in Arm B achieved a median Jaccard index of 0.450 versus 0.259 for the human definitions, median pairwise agreement of 0.492 versus 0.149, and a 5.06-fold versus 32.0-fold size spread. Agent variation resolved almost entirely onto two criteria reported differently in the source main text and supplement. In Arm C, median agreement and median pairwise agreement both reached 1.000.

**Discussion:** Changing only the analyst strongly trended towards improvement in fidelity, with every agent run more self-consistent than every human team. Changing the specification resulted in exact fidelity and self-consistency. Irreproducibility arose from both specification quality and inter-analyst variation.

**Conclusion:** AI agents were more consistent than human teams when translating published natural-language descriptions into code.

## BACKGROUND AND SIGNIFICANCE

Observational studies of administrative health data in real-world settings drive regulatory decisions, coverage policies, and clinical guidelines, yet study samples can vary widely even when drawn from the same source population and analytic cohorts are rarely reproducible from published protocols alone.[1-4] Creating an analytic cohort involves a litany of deliberate decisions and choices: which administrative codes constitute the exposure, how long a defensible washout period is, and whether a diagnosis recorded at an inpatient visit rather than any encounter measures a different construct. Manuscripts report these choices as prose, which rarely captures the full decision trail needed to reconstruct a valid cohort.

Analysts given the same data and hypothesis often reach different conclusions, a pattern documented in psychology,[5] the social sciences,[6] microeconomics,[7] and neuroimaging, where 70 teams produced no two identical pipelines.[8] The REPEAT initiative found substantial variation among studies that attempted to address the same question,[9, 10] and the Observational Health Data Sciences and Informatics (OHDSI) program has invested heavily in phenotype evaluation tooling because clinical definitions tend to vary sub-stantially between studies.[11-14] In a study conducted at an OHDSI symposium, Ostropolets et al. brought together nine expert teams and gave to each the same published study,[15] an identical pre-loaded cohort-building environment with the same concept sets, and eight hours to reproduce the target cohort in the published study.[16] Despite these details provided, the resulting cohorts ranged from 2159 to 63 619 patients against a reference of 6196, and matched the reference on only four of ten criteria on average; chronic lower respiratory disease (CLRD) ascertainment and exclusion for pregnancy were not reproduced as specified by any team. The authors attributed this not to “the skill of the investigators but the process for generating and disseminating evidence itself.”[16] Their conclusion prior to widespread availability of artificial intelligence was: “Natural language description can be generated from the code in an automated fashion. On the other hand, code cannot be generated reliably based on the description, which may be inconsistent, ambiguous, or missing sufficient detail in the paper, diagrams, templates, and Supplementary Materials.”[16]

These tools establish whether a concept definition is sound before implementation, leaving the translation of protocol into executed code manual and analyst-dependent. More recently, AI agents that take over the next steps of writing and executing analytic code have risen in availability. Early evidence raised concerns: left unsupervised to design and implement an observational study from end-to-end, general-purpose LLMs succeeded on fewer than 40% of benchmark tasks[17] and hallucinated medical concept identifiers in 21% to 50% of attempts depending on the model.[18] Beyond task failure, observed nondeterminism, in which identical prompts yield different analyses even at perceived deterministic settings,[19] has been raised as a core barrier to trusting agent-generated evidence.[20] Coding agents have reproduced published social science findings at high rates: given the original data and code, 78% of papers were reproduced in full;[21] given only the methods text, 75 to 91% of the coefficients an agent produced matched the sign of the original, although agents produced only about 81% of the target coefficients.[20] For clinical research, LLMs have been able to reliably draft phenotyping algorithms,[22] parse complex patient information from electronic health record (EHR) systems,[23] and map local schemas to common data models.[24] Nonetheless, whether an agentic AI-assisted workflow can match or surpass the reliability of human analysts has not yet been fully characterized.

This study replicates the OHDSI symposium experiment with the addition of independent AI agent runs in comparison to human analyst teams. We deterministically reproduce the cohorts from the original research investigation[15] and the OHDSI team[16] in HealthVerity claims data from the published code on Medeloop Analytics, an agentic AI research platform. Additionally, we extend the original experiment by giving the agentic AI the same source paper and frozen code sets as the human teams. Because runs across agents can be set to originate from an identical starting state, unlike the interpersonal differences inherent across human teams, repeated runs in Medeloop Analytics allow us to separate what a human experiment could not: how much irreproducibility is attributable to differences in individual analysts, and how much is inherent uncertainty in the reference specification criteria.

## MATERIALS AND METHODS

### Study design

We executed 27 independent reproductions of a published cohort definition across three arms of nine runs each, all against a single database, and scored against a single reference cohort (Figure 1). Arm A reimplemented the nine human team definitions published by Ostropolets et al.[16] as a calibration arm, establishing human performance on our database. Arm B, the primary arm, was nine independent AI agent runs given the source study as published. Arm C was nine further runs given it rewritten to remove its internal contradictions. Hypotheses, arms, and the primary endpoint were fixed before Arms B and C ran.

**Figure 1.**
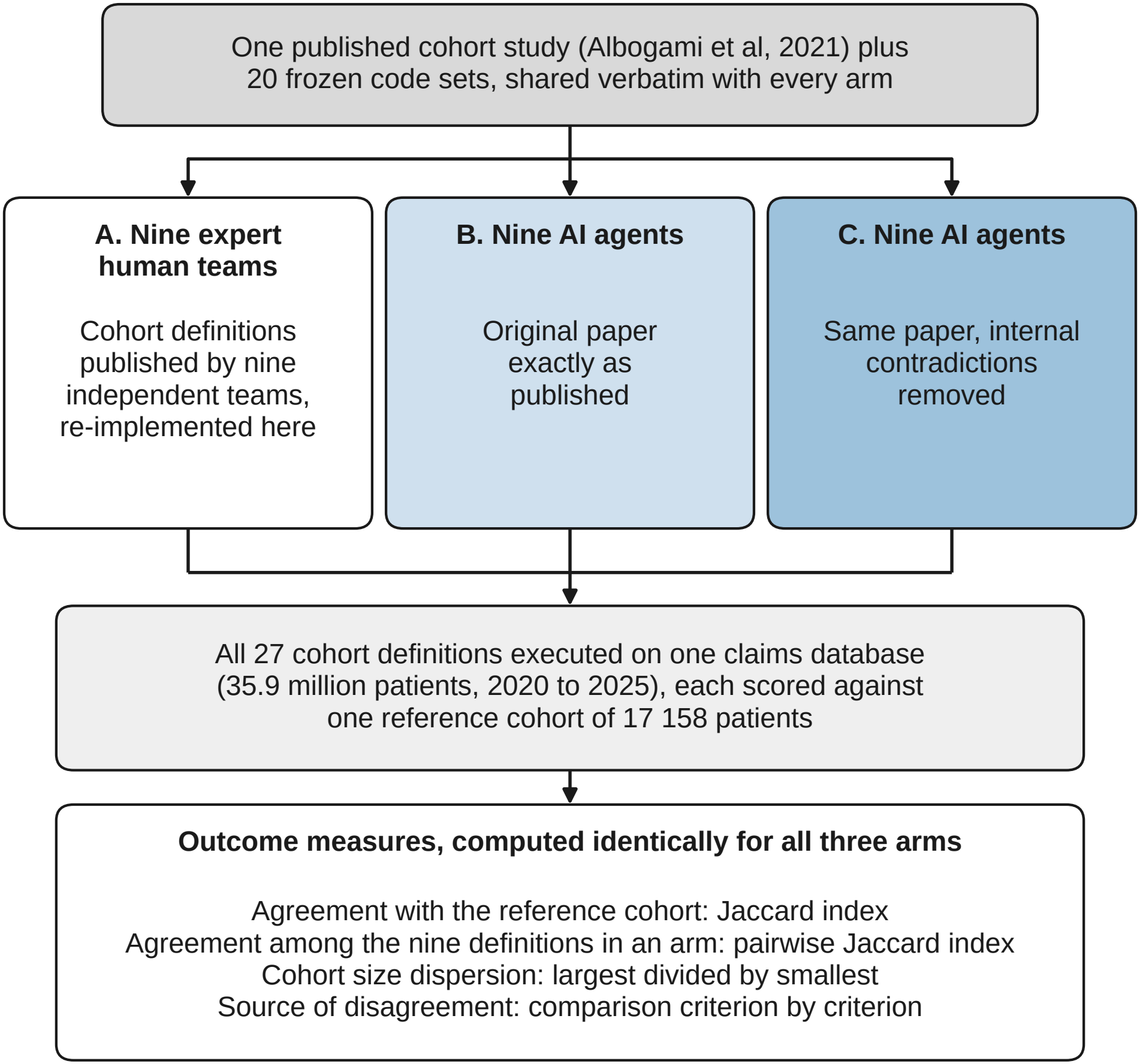
Study design: one published cohort definition, reproduced by three independent arms of nine and scored against a single reference cohort. One published cohort study and a library of 20 frozen code sets were supplied to every arm. Arm A comprised the cohort definitions published by nine expert human teams, re-implemented here. Arm B comprised nine independent artificial intelligence (AI) agent runs given the source paper as published. Arm C comprised nine further runs given the Albogami et al. paper’s methods section rewritten to remove its internal contradictions. All 27 definitions were executed against one administrative claims database of 35.9 million patients spanning 2020 to 2025 and scored against one reference cohort of 17 158 patients on four measures: agreement with the reference cohort; agreement among the nine definitions within an arm; cohort size dispersion; and criterion-level source of disagreement.

### Data source

All 27 definitions ran against HealthVerity Base, a United States administrative claims database of closed medical and pharmacy claims covering 35 874 295 patients as of March 2026. Diagnoses use International Classification of Diseases, Tenth Revision, Clinical Modification codes; dispensings National Drug Codes; procedures Current Procedural Terminology and Healthcare Common Procedure Coding System codes. Because coverage begins in 2020, the index window was mapped to 1 January 2021 through 31 October 2025, preserving the original 365-day lookback and ending before an identified claims run-out period. Relative durations were preserved absolutely; only the absolute window shifted.

### Target study and reference cohort

The target was the new-user glucagon-like peptide-1 receptor agonist (GLP-1RA) arm of Albogami et al.,[15] the study used by Ostropolets et al.[16] The reference cohort was built by porting the original investigators’ intended definition — the specification the nine teams were attempting to recover — from its published machine-generated SQL, yielding 17 158 patients (Supplementary Table S3). This reference is the ground truth for all three arms. We scored against the investigators’ code rather than our own reading of their prose because the prose is the object under test: a reference built by interpreting the paper would encode one particular resolution of the very ambiguities being measured, and every arm would then be scored against an arbitrary choice. The code departs from the published text in several places, including a pregnancy window matching neither the paper’s prose nor its own supplementary figure. “Fidelity” here means fidelity to that intended specification, not clinical correctness.

### Frozen code sets

Both AI arms received an identical library of 20 code set files, and the Arm A re-implementations drew on the same sets. These derive from the concept sets published with the original definitions, which we verified are semantically identical across all ten, so all measured variation is logic, not codes. The library omits the three visit-type concept sets, which this database encodes as columns rather than code lists; the Arm B and C instructions direct agents to those columns.

An additional two of the originally supplied code sets — a CLRD drug list and a corticosteroid-indication list —have no published basis in the target study. They were shared with the human teams through the original cohort-building environment, so they belong in a faithful reconstruction, but the unpublished CLRD drug list plausibly enabled the dominant interpretive fork reported below — an inherent property of the original experiment.

### Arm A: re-implementation of the human definitions

Each of the nine published team definitions was translated into an equivalent query against the claims schema, built from its authoritative compiled SQL. Every translation was reviewed by an independent adversarial reviewer whose remit was to find discrepancies, and revised until review passed. Two cohorts contain criteria whose coded logic is the inverse of the description; we reproduced those cohorts as coded. Construction, review, determinism testing, and the assessment of fidelity against the values published on the original database are detailed in the Supplementary File (Supplementary Table S1).

### Arm B: AI agent runs using the original paper

The AI agents within Medeloop Analytics, an agentic analytic platform, were given schema access and ran query execution against the same database as the re-implementation of the human definitions. Nine runs were conducted in nine separate accounts, each starting from an identical state with no persistent memory and no access to other runs’ conversations, decisions, or outputs. Each received the complete Albogami paper and supplement as PDFs, extracts of the methods, and the frozen code set library.

A single instruction, byte-identical across all nine runs, was issued with no preamble or added context:

“… reproduce the target cohort — the new-user GLP-1 receptor agonist arm — defined in the excerpts below from the Albogami et al. paper and figures 1 & 2 from the supplementary materials … use them as given, do not re-derive or modify them — only the cohort logic is yours to determine.”

The instruction is reproduced verbatim and in full in the Supplementary File. The Ostropolets paper, our reference cohort size, and all intermediate results were withheld.

After each run, the agent’s generated code was read in full and compared criterion by criterion against its own reported attrition and the reference definition, to distinguish interpretive choices from implementation defects. Each analysis was independently reviewed.

### Arm C: AI agent runs from a de-contradicted specification

We rewrote the target study’s methods as continuous prose, removing ambiguity and resolving every internal contradiction to a single reading based on the published SQL code. Nine runs, conducted under the same isolation in nine additional unique workspaces, received this prose as instruction. Because the specification describes the reference cohort as built, Arm C measures convergence on a described target, not recovery of an undisclosed one. The rewrite names no database, discloses no target count; like Arm B’s instruction, it names the source paper but not this study. Two count-free companion figures were redrawn to match the supplementary figures from Albogami et al. (Supplementary Figures S2 and S3).

### Outcomes and analysis

The primary endpoint was fidelity to the investigators’ intended definition, recreated in HealthVerity Base: the Jaccard index between each cohort and the reference, the number of patients in both divided by the number in either. The secondary endpoint was reproducibility: the pairwise Jaccard index between units within an arm. Other measures include the median Jaccard index across all 36 within-arm pairs, cohort-size dispersion, and a criterion-level comparison classifying each definition’s reading of ten shared criteria against the reference. All tests were non-parametric, both because the outcomes are bounded proportions and because Arm C has zero within-arm variance (every run identical), which precludes parametric methods.

For fidelity (nine values per arm), we used the Kruskal–Wallis test for the overall difference among arms, the Jonckheere–Terpstra test (permutation-based) for the pre-specified ordered alternative A < B < C, and pairwise two-sided Mann–Whitney U tests with Holm correction for the three arm contrasts; effect sizes are reported as Cliff’s delta. For reproducibility, the run — not the pair — is the unit of analysis, because the 36 within-arm pairwise Jaccard indices are not independent (each run appears in eight pairs). Between-arm reproducibility was therefore tested with a label-permutation test that permuted run-to-arm assignments and recomputed each arm’s within-group median pairwise Jaccard (50 000 permutations, two-sided), complemented by a unit-level Mann–Whitney U test on each run’s mean pairwise Jaccard with Cliff’s delta. Analyses were performed in Python (SciPy).

### Disclosure of AI use

This study evaluates an AI system, and AI was additionally used in conducting and reporting it. Medeloop Analytics was the object of study in Arms B and C. Anthropic Claude models were used to build and review the Arm A re-implementations, translate the specification for Arm C, compute the comparisons, generate the figures, and suggest edits to this manuscript. All quantitative results were verified against archived run outputs, and all authors accept responsibility for the content. No AI system is listed as an author.

## RESULTS

### Human team definitions reproduce poorly on a common database

Re-executing the nine published human definitions produced cohorts of 2614 to 83 706 patients against a reference of 17 158 — a 32.0-fold spread (Table 1, Table 2, Figure 2A). The median Jaccard index against the reference was 0.259 (range 0.000–0.508). Median pairwise agreement between team definitions was 0.149; eight of 36 pairs shared no patients at all. Two definitions were near-disjoint from the reference — Team 4 at 0.013, Team 5 at 0.000 — because criteria labelled insulin exclusions are coded as insulin requirements, selecting nearly complementary populations. Agreement ranks correlate with the values published on the original database (Spearman ρ = 0.946), as do per-team cohort sizes (ρ = 0.967), and both extremes are preserved, the basis for treating this arm as a faithful calibration (Supplementary Table S2; Supplementary Figure S1).

**Table 1.** Cohort size and agreement with the reference cohort across three reproduction arms.

|  | **Human team definitions** | **AI agents, original paper** | **AI agents, revised specification** |
|---|---|---|---|
| Reproductions, n | 9 | 9 | 9 |
| Cohort size, median | 31 381 | 14 762 | 17 158 |
| Cohort size, range | 2614–83 706 | 13 269–67 115 | N/A |
| Size dispersion | 32.0-fold | 5.06-fold | N/A |
| Jaccard vs reference, median | 0.259 | 0.450 | 1.000 |
| Jaccard vs reference, range | 0.000–0.508 | 0.222–0.490 | N/A |
| Pairwise Jaccard, median | 0.149 | 0.492 | 1.000 |
| Pairwise Jaccard, range | 0.000–0.843 | 0.189–0.976 | N/A |
| Near-disjoint cohorts, n | 2 (0.013, 0.000) | 0 | 0 |
| Patient-identical clusters | 0 | 0 | 1 |

Reference cohort n = 17 158. All arms executed against the same database with identical frozen code sets and an identical index window; only cohort logic varied. The Jaccard index is the number of patients in both cohorts divided by the number in either. Pairwise values summarise all 36 within-arm pairs. “Near-disjoint” denotes a Jaccard index below 0.05 against the reference cohort.
Abbreviation: AI, artificial intelligence.

**Table 2.** Individual reproductions, by arm.

|  | **Arm A: human team definitions** |  | **Arm B: AI, original paper** |  | **Arm C: AI, revised specification** |  |
|---|---|---|---|---|---|---|
|  | **n** | **Jaccard** | **n** | **Jaccard** | **n** | **Jaccard** |
| 1 | 73 560 | 0.210 | 13 509 | 0.461 | 17 158 | 1.000 |
| 2 | 41 020 | 0.266 | 66 824 | 0.222 | 17 158 | 1.000 |
| 3 | 14 886 | 0.259 | 14 915 | 0.490 | 17 158 | 1.000 |
| 4 | 17 995 | 0.013 | 67 115 | 0.222 | 17 158 | 1.000 |
| 5 | 2614 | 0.000 | 13 879 | 0.426 | 17 158 | 1.000 |
| 6 | 9155 | 0.285 | 13 269 | 0.450 | 17 158 | 1.000 |
| 7 | 83 706 | 0.189 | 14 762 | 0.481 | 17 158 | 1.000 |
| 8 | 31 381 | 0.508 | 13 809 | 0.468 | 17 158 | 1.000 |
| 9 | 45 329 | 0.300 | 52 056 | 0.271 | 17 158 | 1.000 |
| Median | 31 381 | 0.259 | 14 762 | 0.450 | 17 158 | 1.000 |

Reference cohort n = 17 158. Row numbers index independent reproductions within an arm and are not comparable across arms.
Arm A rows correspond to the numbered teams of the source reproduction study; Arm B and Arm C rows are separate agent runs.
Abbreviation: AI, artificial intelligence.

**Figure 2.**
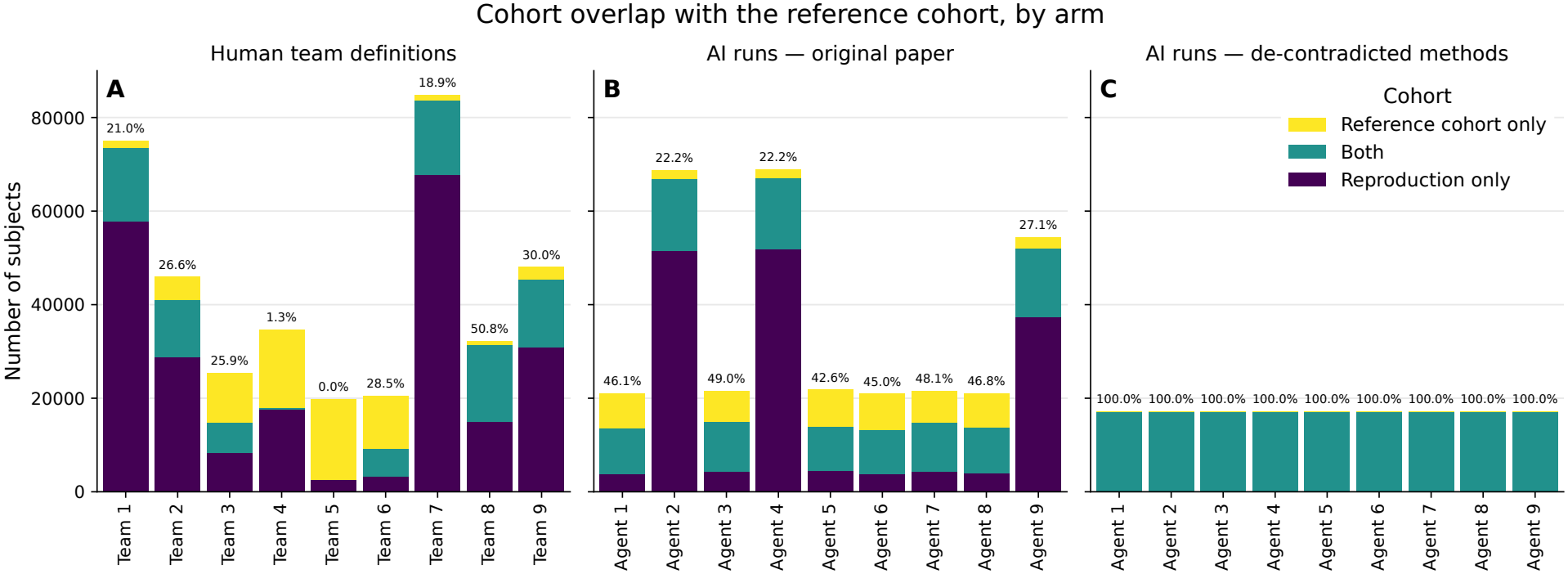
Cohort overlap with the reference cohort, by arm. Nine expert human teams and nine artificial intelligence (AI) agents working from the same published paper both produced cohorts that overlapped the reference cohort only partially; nine agents given the same methods with their internal contradictions removed reproduced the reference cohort exactly. Each bar is one reproduction of the same published glucagon-like peptide-1 receptor agonist cohort definition and spans every patient in either that reproduction’s cohort or the reference cohort. Within each bar, the lower segment is patients selected by the reproduction only, the middle segment patients in both, and the upper segment patients in the reference cohort only, so the lower two sum to the reproduction’s cohort size and the upper two to the reference cohort size of 17 158. The value above each bar is that reproduction’s Jaccard index against the reference cohort as a percentage — patients in both cohorts divided by patients in either — so 100% denotes identity and 0% no shared patients. (A) The nine human team definitions re-implemented here; agreement 0.0% to 50.8%, median 25.9%, with two teams sharing essentially no patients with the reference (Team 4, 1.3%; Team 5, 0.0%). (B) Nine independent AI agent runs given the source paper as published; agreement 22.2% to 49.0%, median 45.0%, the cohorts falling into two size groups differing roughly fourfold. (C) Nine further runs given the same methods rewritten to remove their internal contradictions; every run returned the reference cohort exactly, each bar consisting of the shared segment alone. Segments differ in lightness as well as hue.

### AI agents given the same source outperformed the human teams

Nine independent agent runs, given the same paper and code sets, produced cohorts of 13 269 to 67 115 patients — a 5.06-fold spread against the human 32.0-fold. Median agreement with the reference was 0.450 (mean 0.388, range 0.222–0.490), against the human median of 0.259 (Table 1, Figure 3, left). Seven of nine exceeded the human median; the two that did not scored 0.222, still far above the two lowest scoring human definitions of 0.013 and 0.000.

**Figure 3.**
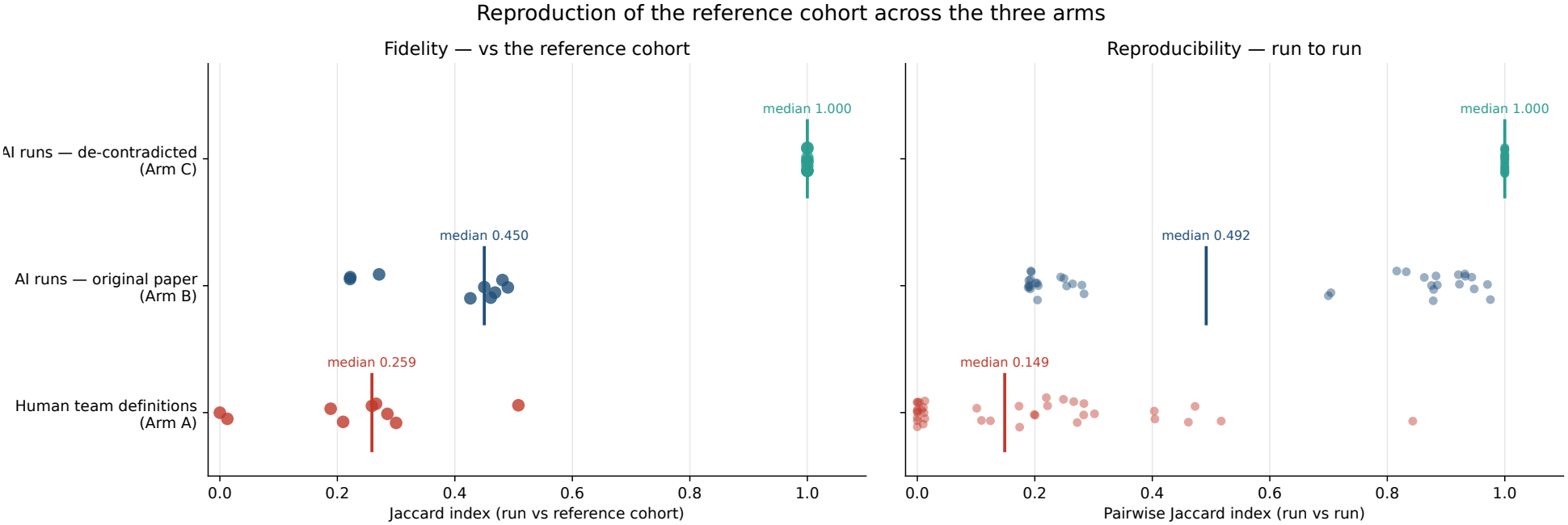
Fidelity and run-to-run reproducibility across the three arms. Removing the source paper’s internal contradictions moved nine independent AI agent runs from partial agreement to exact reproduction, on both fidelity to the reference cohort and agreement with one another. Each point is one measurement and the vertical bar marks the arm’s median; points are jittered vertically for visibility only. The Jaccard index is defined as in Figure 2, here on a 0–1 scale. Left, fidelity: nine points per arm, one per reproduction, against the reference cohort; medians were 0.259 for the re-implemented human team definitions, 0.450 for the AI runs given the original paper, and 1.000 for the AI runs given the de-contradicted methods. Right, reproducibility: 36 points per arm, one per within-arm pair; because each reproduction appears in eight pairs these points are not independent, and the arm is the unit of inference. Medians were 0.149, 0.492 and 1.000 respectively. In the third arm every value is exactly 1.000, so the apparent vertical spread is jitter alone. The bimodality in the second arm’s pairwise values reflects two groups of runs that read one contradictory rule in opposite ways; see Figure 4.

Consistency separated the arms further: median pairwise agreement among agent runs was 0.492, against 0.149 among human definitions. Both pre-specified hypotheses were supported. Fidelity differed across the three arms (Kruskal–Wallis P < .001) in the pre-specified order A < B < C (Jonckheere–Terpstra P < .001). Holm-corrected pairwise contrasts gave P = .064 for Arm A versus Arm B (Cliff’s δ = +0.53) and P < .001 for Arm C against each of the other two arms (δ = 1.00). Reproducibility separated the arms completely: every run was more self-consistent than every run in the arm below it (run-level Mann–Whitney *P* < .001 for each contrast; Cliff’s δ = +1.00). Permuting run-to-arm labels gave the same conclusion for the human-versus-agent contrast (*P* < .001).

### Agent variation was structured, and localised to two contradictory passages

Human deviation was diffuse: teams in Arm A differed from the reference on three to seven criteria each, spread across the definition. Agent variation in Arm B was not. It resolved almost entirely onto two passages where the source paper contradicts itself (Figure 4).

**Figure 4.**
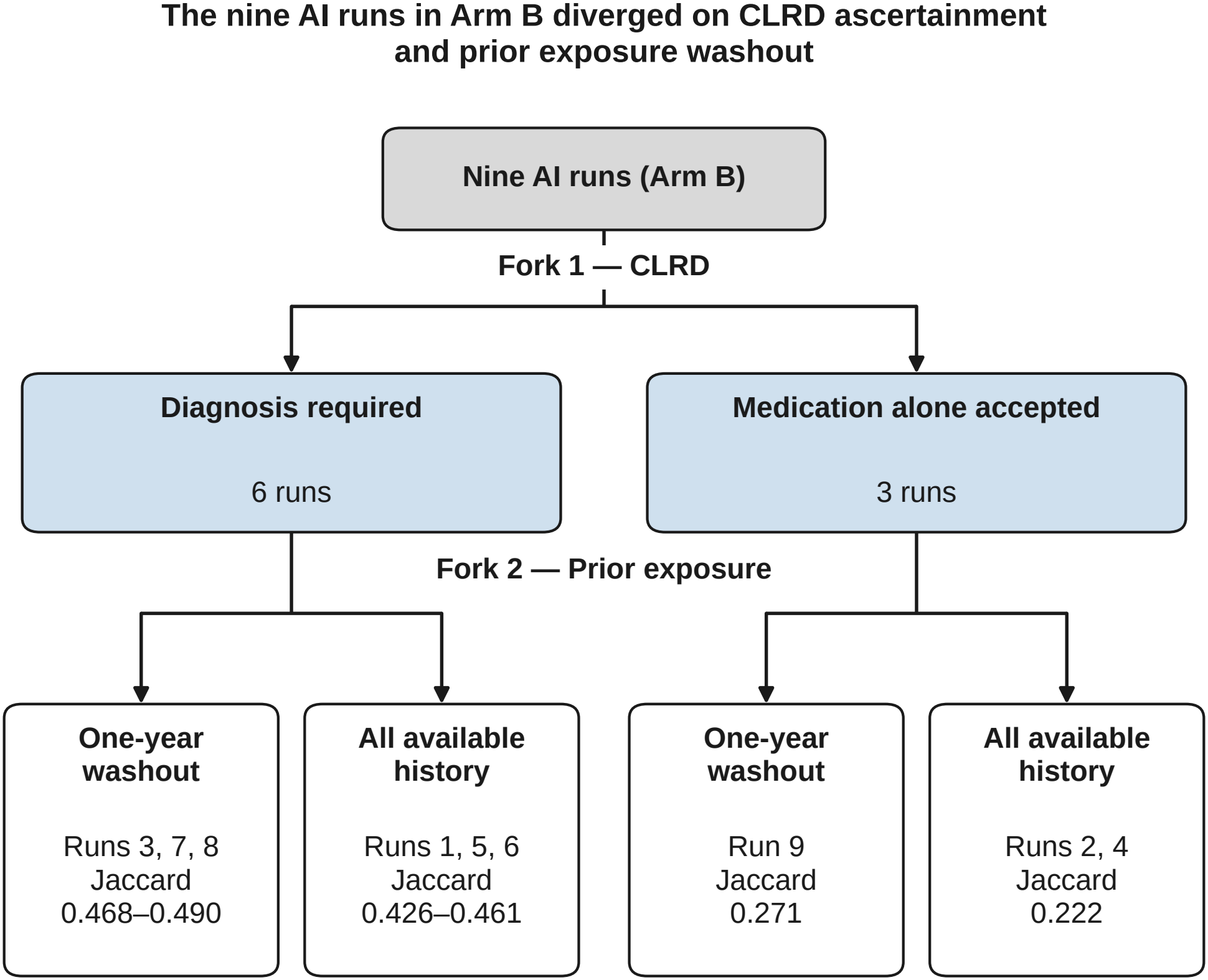
The nine AI runs in Arm B diverged on CLRD ascertainment and prior exposure washout. A decision tree classifying the nine independent artificial intelligence (AI) agent runs given the source paper as published, by the two interpretive choices that separated them; each terminal box names its member runs and the range of their Jaccard indices against the reference cohort. The first split is ascertainment of chronic lower respiratory disease (CLRD), on which the source paper’s prose admits a qualifying medication alone while its own supplementary table and attrition figure describe a diagnosis requirement: six runs required a diagnosis and three accepted a medication. The second is the prior-exposure washout, on which the prose implies all available patient history while a supplementary figure depicts a one-year window: four runs used one year and five used all history. Agreement ordered monotonically across the four terminal boxes, from 0.468–0.490 where a diagnosis was required with a one-year washout, through 0.426–0.461 for a diagnosis with all history, to 0.271 for a medication with a one-year washout and 0.222 for a medication with all history. Abbreviation: CLRD, chronic lower respiratory disease.

The dominant axis was CLRD ascertainment: the paper’s prose admits a qualifying CLRD *medication* alone, while its own supplementary table and attrition figure describe a *diagnosis* requirement. Six runs required a diagnosis; three accepted a medication. This single choice moved cohort size by roughly a factor of four and agreement by approximately 0.22. The secondary axis was the prior-exposure washout: the prose frames new use as the first dispensing after 365 days of continuous enrollment, not as a lookback, while a supplementary figure depicts a one-year window. Four agents used one year, five used all history, with only a small impact to the pairwise Jaccard index of 0.03–0.06.

The two axes define four cells that order agreement monotonically (Figure 4), from 0.468–0.490 for diagnosis with a one-year washout down to 0.222 for medication with all history. Median pairwise agreement was 0.885 among the six diagnosis-requiring runs but 0.193 across the CLRD divide (Figure 3, right).

Both readings are defensible and the intended resolution was unknowable: the investigators’ own diagram of the reference definition included in Ostropolets et al., which the human teams never saw, silently resolves both forks and implements a third, hybrid CLRD rule.

### Under an unambiguous specification the agents converged

In Arm B, there were two major decision forks that resulted in clusters (Figure 4). In Arm C, nine fresh runs given the methods rewritten to a single consistent reading produced cohorts of exactly 17 158 patients — identical to the reference cohort (Figure 3). Both fidelity and pairwise agreement were 1.000.

### Disagreement tracked specification quality, not criterion difficulty

Classifying every definition’s reading of ten shared criteria against the reference reveals the pattern (Table 3). On the four criteria the source specifies inconsistently, deviation was near-universal in both Arm A and Arm B: CLRD 9 of 9 in both, pregnancy 9 of 9 in both. On the two the source specifies unambiguously, deviation was near-absent in both: no definition in either arm differed on age, and one in each differed on the other medical exclusions. Human definitions differed on 50 criterion readings in total (Arm A), while agent runs (Arm B) differed on 62 criterion readings, of which 61 relied on criteria specified incompletely or in a different manner across study materials; the remaining deviation was a single run that excluded type 1 diabetes over all available history rather than the specified baseline year.

**Table 3.** Interpretation of ten shared criteria, by how the source paper specifies each.

| Criterion | Specification in source | Human definitions deviating (Arm A), n/9 | AI runs deviating (Arm B), n/9 |
|---|---|---|---|
| Chronic lower respiratory disease | Inconsistent | 9 | 9 |
| Type 2 diabetes ascertainment | Inconsistent | 8 | 9 |
| New-user exposure washout | Inconsistent | 7 | 7 |
| Pregnancy exclusion | Inconsistent | 9 | 9 |
| Continuous enrollment, 365 days | Ambiguous | 0 | 9 |
| Add-on therapy requirement | Ambiguous | 8 | 9 |
| Insulin washout | Ambiguous | 3 | 4 |
| Dipeptidyl peptidase-4 inhibitor washout | Ambiguous | 5 | 5 |
| Other medical exclusions | Unambiguous | 1 | 1 |
| Age restriction | Unambiguous | 0 | 0 |
| Total criterion deviations |  | 50 | 62 |

“Deviating” means the reproduction’s reading of that criterion differs from the reference definition’s, irrespective of the difference’s effect on the resulting cohort. Criteria are classified by whether the source study states them once and consistently (unambiguous), once but incompletely (ambiguous), or more than once with conflicting content across text, tables, and figures (inconsistent). Human readings were taken from the source reproduction study’s published deviation table and verified against our re-implementations. Our count of 50 differs by one from the 51 enumerated in that study’s own figure; the discrepancy is confined to a single insulin-washout cell on which the study’s table and figure disagree. The higher agent total is driven by continuous enrollment and add-on therapy, on which all nine runs adopted the same near-inert looser reading; see Results.
Abbreviation: AI, artificial intelligence.

That the agents in Arm B differ on *more* criteria while agreeing far better on *patients* is not a contradiction. Ten of the twelve extra agent deviations fall on two near-inert criteria — continuous enrollment (nine) and add-on therapy (one) — where all nine runs adopted the same slightly looser reading, attributable to this database’s enrollment representation differing from the common data model the original definitions assumed.

## DISCUSSION

Given the same published description, code sets, and database, an AI agent reproduced a cohort definition roughly 1.7 times more faithfully than nine expert human teams (median Jaccard 0.450 versus 0.259) and 3.3 times more consistently (pairwise 0.492 versus 0.149), with no near-disjoint cohorts resulting, compared with two among the human team runs. Reconciling criteria reported differently by the source across its main text and supplement raised the mutual agreement among nine fresh runs (Arm C) to 1.000.

Our result refines the claim from Ostropolets et al. that code cannot be reliably generated from description because it “may be inconsistent, ambiguous, or missing sufficient detail.”[16] Our findings support that mechanism and add both magnitude and a remedy. The same platform, on the same database with the same code sets, moved from a split two ways on a self-contradictory sentence (Arm B) to one whose independent runs returned identical patients (Arm C), with no change to the AI analyst. However, a qualification is that the rewrite of the source methods (Arm C) was authored with knowledge of the code executed by Albogami et al., so it shows an unambiguous specification *can* be written, not that authors will write one unaided.

The design separates the two sources of variation. Holding the source description constant and changing only the analyst — Arm A to Arm B — left fidelity statistically unchanged: the median Jaccard index rose from 0.259 to 0.450, a large but non-significant effect (Cliff’s δ = +0.53; P = .064). The same change separated the arms completely on consistency, every agent run being more self-consistent than every human team (run-level Mann–Whitney P < .001; δ = +1.00). Holding the analyst constant and changing only the specification Arm B to Arm C — moved both to exact (Holm-adjusted P < .001; δ = 1.00). The analyst therefore accounts for how consistently a cohort definition is rebuilt; the specification accounts for how accurately it is rebuilt, and only the second is remediable by better reporting.

### Scope, and comparison with contemporaneous benchmarks

These results characterise one commercial platform on one task, not a property of AI agents in general. Three contemporaneous evaluations report far weaker performance, and those differences are instructive rather than contradictory.

RWE-bench evaluated six models across three agent scaffolds on 162 observational-study tasks drawn from an intensive-care database; the best configuration succeeded on 39.9%, and the same model varied from 1% to 30% across scaffolds.[17] Neither the task nor the metric matches ours. Their tasks run through to effect estimation, and success is conjunctive — every question and every statistical field must be correct.

A third reproduction of the same target study differs from the other two in its input rather than its analyst: Conover et al. worked from the original analytical code, furnished by the Albogami investigators, and recov-ered the cohorts to near-identity, with every confidence interval containing the original point estimates.[25] Set beside the nine teams who had only the published prose, it is consistent with the remedy Ostropolets et al. proposed — that code, not better analysts, is what carries a cohort definition intact. Our numbers are nonetheless conditional on the codes being given. Lee et al. converted free-text eligibility criteria to common-data-model queries *without* frozen code sets and found concept identifiers hallucinated in 32.7% of 760 query-generation attempts overall, with Jaccard indices against reference definitions of 0.00, 0.03, 0.18 and 0.81.[18]

RWE-bench attributes part of its agent failures to ambiguous cohort criteria producing divergent inclusion decisions, and recommends requiring agents to expose granular decision-making.[17] Cui and Alexander, running 480 analyses across six models and four temperatures, found run-to-run variation persisting even at temperature zero.[26] Specification ambiguity and run-level dispersion are reproducible findings and not the result of any one platform.

### On non-determinism in agentic AI

Nondeterminism is not unique to LLMs, but it is easily identifiable in them. Human analysts given identical variable definitions and protocols struggle to produce concordant cohorts.[16] Agents can easily and quickly rerun analyses from identical starting points at marginal cost, making reproducibility across runs quantifiable and reportable. Repeated runs make the problem observable, with divergence across runs marking the decisions a protocol left open. The practical requirement for agentic workflows is therefore not determinism, but what these systems inherently provide: measurable variability and full traceability at every step.

### Limitations

Comparisons between human teams and agents warrant several considerations. The human teams worked within eight hours; our runs had no time limit. Nonetheless, arm mean run times ranged from 15.8 to 43.5 minutes (Supplementary Table S4). Human teams consisted of experienced investigators who could deliberate and apply clinical judgement, but carried analytic preferences informed by their own training and practice that may have influenced interpretations of the source text; likewise, the agents’ decisions were subject to the same risk from their training. The human definitions are a fixed historical artefact, and because we could not access the original database, we cannot separate how much of our uniformly higher agreement (0.259 against the 0.094 that study reports, or 0.153 recomputed as the median of its nine published per-team values; Supplementary Table S2) reflects the different population from how much reflects the claims schema.

We evaluated one platform, without changes to the agents, the underlying LLM models, or the temperature. Cui and Alexander recommend sampling models from at least two providers before drawing conclusions about LLM-assisted analysis;[26] the consistency we report may well be a property of this configuration rather than of agentic analysis in general. The 2023 Ostropolets et al. publication and its openly available cohort definitions may be in the LLM training data. Two observations mitigate this: the reference cohort was materialised on a database the original investigators never used, so no published count corresponds to our target; the *pattern* of Arm B’s split — two stable clusters straddling one contradictory sentence — is produced by interpretation rather than recall. Further limitations: the Jaccard index measures membership overlap only and is blind to covariate balance, which the original study also assessed; the database begins in 2020, left-censoring prevalent users at the window’s start. Fidelity means fidelity to the intended specification. Finally, this is a reproduction of a reproduction study, on one cohort, in one therapeutic area.

### Implications

For authors and journals, the actionable target is the specification. Both criteria that split the agents in Arm B were ones the source stated more than once and differently; the two criteria that were stated once and consistently produced near-complete agreement. Presenting criteria across text, tables, and figures is standard practice, yet few reporting guidelines require reconciliation or the release of executable code for direct verification. Internal consistency is low-cost and mechanically checkable. Executable definitions remain the stronger remedy,[27, 28] but consistency is available to every paper immediately.

For evaluating AI analysts, single runs are uninformative. Our nine Arm B runs spanned 13 269 to 67 115 patients; any one, reported alone, would have looked like a clean result. Reporting *where* runs diverge is more informative still: the pattern of divergence located a defect in the source paper that reading it had not surfaced. A set of independent runs disagreeing in a structured way is an ambiguity detector.

## CONCLUSION

Given the same source description and code sets, an AI analytic agent reproduced a published cohort definition more faithfully and far more consistently than nine expert human teams, producing no cohort essentially disjoint from the reference, whereas two human definitions did. Its remaining disagreement was concentrated on the two passages where the source paper contradicted itself; under a specification without those contradictions, independent runs returned patient-identical cohorts. For this platform and this task, description-to-code generation is more reliable than previously concluded — provided the description is internally consistent.

## REQUIRED STATEMENTS

## Data Availability

The claims database analysed here (HealthVerity Base) is licensed from HealthVerity and cannot be redistributed by the authors; access is available to third parties directly from HealthVerity under commercial licence. No patient-level data left the analytic environment at any point, and none is available for sharing.
The published cohort definitions reproduced in Arm A are openly available from the original investigators at https://github.com/aostropolets/ReproducibilityChallenge. Derived artefacts generated by this study are available at https://github.com/medeloop/public: the ten deterministic re-implementations of the published cohort definitions; the 20 frozen code sets, with a checksum script; the pairwise agreement matrices for all 27 reproductions, with the script that recomputes every statistic reported here from them; all aggregate cohort counts, agreement indices and run times reported in this article; and the figure generation scripts with their tabulated source data, which regenerate all four main-text figures and Supplementary Figures S1 to S3. All analyses reported in this article are repeatable from the deposited aggregate data without access to patient-level records.

## Acknowledgements

We thank the investigators of both source studies for publishing their cohort definitions as executable code and for making the nine team definitions openly available; this study would not have been possible otherwise.

Generative AI was used in this work; see “Disclosure of AI use” in Materials and Methods for the full statement.

## Author contributions

JW: Conceptualization; Data curation; Formal analysis; Investigation; Methodology; Project administration; Resources; Software; Supervision; Validation; Visualization; Writing – original draft; Writing – review & editing.

CE: Conceptualization; Methodology; Software; Validation; Writing – original draft; Writing – review & editing.

AJ: Conceptualization; Methodology; Software; Writing – review & editing.

AB: Data curation; Software.

MF: Software.

YC: Software; Writing – review & editing.

## Competing interests

The AI system evaluated in Arms B and C, Medeloop Analytics, is a commercial product of Medeloop, of which all authors are employees and in which all hold an equity stake. The authors therefore have a direct financial interest in the finding that this system performs well. Readers should weigh the results accordingly.

To limit favourable bias we fixed the following before Arms B and C were executed: the hypotheses and primary endpoint; the reference cohort, published here with its cohort hash; and the byte-identical run instruction. Every Arm A re-implementation was subjected to independent adversarial review whose explicit remit was to find discrepancies.

We did not pre-register publicly, employ a blinded outcome assessor, or have a non-conflicted party execute the runs.

## Ethics approval

This study uses data that are expert-certified as de-identified under HIPAA. No IRB approval was required.

## Data availability statement

The claims database analysed here (HealthVerity Base) is licensed from HealthVerity and cannot be redistributed by the authors; access is available to third parties directly from HealthVerity under commercial licence. No patient-level data left the analytic environment at any point, and none is available for sharing.

The published cohort definitions reproduced in Arm A are openly available from the original investigators at https://github.com/aostropolets/ReproducibilityChallenge. Derived artefacts generated by this study are available at https://github.com/medeloop/public: the ten deterministic re-implementations of the published cohort definitions; the 20 frozen code sets, with a checksum script; the pairwise agreement matrices for all 27 reproductions, with the script that recomputes every statistic reported here from them; all aggregate cohort counts, agreement indices and run times reported in this article; and the figure generation scripts with their tabulated source data, which regenerate all four main-text figures and Supplementary Figures S1 to S3. All analyses reported in this article are repeatable from the deposited aggregate data without access to patient-level records.

