## Supplementary materials for "Reproducible variability: assessing human vs. AI discordance in attempting to reproduce the same observational study"

Supplementary material accompanying the manuscript submitted to the *Journal of the American Medical Informatics Association*.

**Contents**

Supplementary Methods 1 — Arm A: construction and validation of the human-definition re-implementations

Supplementary Table S1 — Logic fingerprints, cohort sizes, and cohort hashes

Supplementary Table S2 — Agreement with the reference definition, as published and as obtained here

Supplementary Figure S1 — The same nine team definitions, as published and as re-implemented here

Supplementary Methods 2 — Agent instruction (Arm B)

Supplementary Methods 3 — Input bundle and account isolation

Supplementary Methods 4 — Revised specification (Arm C)

Supplementary Figure S2 — Study design and timeline of baseline assessment windows, as supplied to Arm C

Supplementary Figure S3 — Cohort selection, as supplied to Arm C

Supplementary Table S3 — Reference cohort attrition

Supplementary Table S4 — Run time (minutes) per reproduction, by arm

### SUPPLEMENTARY METHODS 1 — ARM A: CONSTRUCTION AND VALIDATION OF THE HUMAN-DEFINITION RE-IMPLEMENTATIONS

Arm A re-implements, on this study's database, the nine cohort definitions published by the nine expert teams of the reproduction study, together with the definition those teams were attempting to recover, termed the reference implementation in that study. That re-implementation is this study's reference cohort, and the ground truth against which all 27 reproductions in all three arms are scored. This section documents how the ten re-implementations were built, how they were reviewed, how they were shown to be deterministic, and how their fidelity was assessed against the values the reproduction study published on its own database.

#### Source of truth and translation

Every re-implementation was built from the two authoritative artefacts published for that definition: its machine-readable cohort specification, and the SQL compiled from that specification by the cohort-definition compiler. Neither was paraphrased. No re-implementation was built from a flattened or summarised intermediate — not from the reproduction study's prose descriptions of the team definitions, not from its supplementary deviation table, and not from any parse of a specification into a simplified rule list. Where those secondary sources and the compiled SQL disagree, the compiled SQL governs, because the compiled SQL is what the team's definition actually executes. Each implemented rule carries, in its re-implementation's header, the line range of the compiled SQL statement it was translated from, so that every predicate is traceable to the statement that produced it.

Translation was faithful rather than corrective. Every re-implementation carries a standing instruction to execute exactly as written, with no modification, optimisation, correction, or re-ordering, and idiosyncratic or erroneous design choices were reproduced verbatim rather than repaired. Two examples show what this means in practice. Team 8's insulin-exposure exclusion carries a 356-day bound where 365 was plainly intended; the 356-day bound was preserved, and on this database it is provably inert, because the rule's two arms — insulin as a drug and insulin as a device — both resolve to the same dispensing record here and collapse to their union window. Team 8's rule *names* also assert an inpatient-or-two-outpatient encounter requirement for type 2 diabetes that its compiled SQL does not implement; the SQL requires only a diagnosis anywhere in the record, and the rule was implemented as compiled rather than as named. Team 8 is the highest-agreeing team in this study and in the original, and this permissiveness is why. The two definitions whose insulin criteria invert their labels are treated separately below.

Two rules were omitted only after being shown to be inert, never approximated. Team 6's definition includes drug-at-visit arms that are provably empty on this database, because pharmacy dispensings here carry a pharmacy-namespace visit identifier and are therefore never linked to an office, ambulatory-surgery, hospital, or emergency visit; a direct probe returned zero such linkages. Omitting these arms is exact rather than approximate, and the same arms are inert in the common data model the original definitions assumed, where dispensings likewise carry the pharmacy visit identifier. Team 9's definition includes an additional-criteria arm requiring at least one antidiabetes drug era at index, which has no impact because the GLP-1RA code set is a subset of the antidiabetes code set — none of the 172 GLP-1RA product codes falls outside it — so the index dispensing always satisfies it.

One implementation decision was pre-declared and probe-tested rather than assumed. Continuous coverage was required as at least 365 days within a single uninterrupted enrollment span on the medical and, separately, the pharmacy benefit. The alternative — coalescing abutting and overlapping spans, which is how the common data model's observation period behaves — was tested against it with a pre-declared decision rule: adopt the coalesced gate if more than a few per cent of patients pass it while failing the single-span gate. The measured fraction was 0.07%, so the single-span gate was retained for every re-implementation.

##### Independent adversarial review

Every re-implementation was subjected to independent adversarial code review before it was locked. The reviewer's remit was explicitly to find discrepancies between the re-implementation and its source specification and compiled SQL, rule by rule. A re-implementation was not locked until review passed; where review found a discrepancy, the re-implementation was revised and re-reviewed.

Two review-gated revisions, both recorded in the header of the affected re-implementation, illustrate the gate. In the reference definition, review established that the index date must be the earliest in-window dispensing that *itself* satisfies the 365-day coverage requirement, because the compiled SQL applies its first-occurrence operator after the observation filter rather than before it; and that every criterion must be bounded to the mapped absolute window, which caps the forward half of the pregnancy window. Both were corrected before locking. In Team 1's re-implementation, review restored the type 2 diabetes and CLRD criteria to the full three-armed "any of" form after a draft had implemented the drug arm alone, and established that this team's index event is the first-ever qualifying dispensing with no fallback.

#### Logic fingerprints and cohort hashes

Every re-implementation emits two digests.

A **logic fingerprint** is a 16-character truncation of a SHA-256 digest computed over the database identifier, the date configuration, the absolute window bounds, the four SQL predicate tokens that define coverage and care setting, the checksum of every code set the definition uses, and the name and full SQL text of every step. It therefore changes if any input, any window, any code set, or any line of executed logic changes. Two re-implementations sharing a fingerprint would be executing identical logic against identical inputs. Each fingerprint was locked when its re-implementation passed review; it is printed on load, at every step, and at finalisation, and was re-verified on every subsequent run. All ten ran to completion with every fingerprint matching its locked value.

A **cohort hash** is a SHA-256 digest computed over the comma-joined, sorted list of final patient identifiers. Two cohorts sharing a cohort hash are the same set of patients. A per-step digest is emitted alongside it, so any divergence can be localised to the step that produced it.

Per-step patient identifier sets were written to storage inside the account and were never printed; only aggregate counts, fingerprints, and hashes reached the run log. All overlaps between cohorts were computed inside a single account, on patient identifiers that never left it.

**Table S1. Logic fingerprints, cohort sizes, and cohort hashes for the reference definition and the nine Arm A re-implementations**

| Cohort | Logic fingerprint[a] | Cohort n | Cohort hash[b] |
|---|---|---|---|
| Reference | 948bc3640b726cda | 17 158 | e7ae2cdfde68d632…ed59660 |
| Team 1 | 51c7cbe25e23c768 | 73 560 | 61b8d68338ec514b…e7dd978 |
| Team 2 | 6532ae8a04157b74 | 41 020 | 9f8c99f6fb10b3eb…202643c |
| Team 3 | 64b9dbf8dfc6af32 | 14 886 | 29ccc5ec8de16e0b…5fac549 |
| Team 4 | 9feeab7f76aeb32a | 17 995 | ca7e276605f4a713…8ada5ec |
| Team 5 | ab44448b08108b7e | 2614 | 931dfec25d0cbe2d…a8772b5 |
| Team 6 | 03ba0bb93036f925 | 9155 | 1c21a323bacdf8b8…ff37b51 |
| Team 7 | 376e1de53812563f | 83 706 | 3a0e909687f764df…95a8916 |
| Team 8 | b36cbe45a5d1dd84 | 31 381 | 280c8e006cf9fb02…893e36a |
| Team 9 | 775ebe3b81467bb5 | 45 329 | c391c6bf9a0e2ac6…30b13e9 |

All ten re-implementations ran to completion and every logic fingerprint matched its locked value. The values shown are those of the consolidated run, in which all ten cohorts were executed co-located in a single account so that overlaps could be computed on patient identifiers that never left that account.

[a] A 16-character truncation of a SHA-256 digest computed over the database identifier, the date configuration, the absolute window bounds, the four SQL predicate tokens, the checksum of every code set used, and the name and full SQL text of every step.

[b] A SHA-256 digest computed over the comma-joined, sorted list of final patient identifiers. Hashes are shown truncated to their first 16 and last 7 characters; re-running a deposited implementation prints the full

digest.

### Determinism testing

Determinism was tested by executing the same re-implementation in two different accounts and comparing cohort hashes.

All ten cohort definitions produced **byte-identical cohort hashes in both settings**: the same patient set, recomputed in a second account with an independent workspace.

Because all twelve of the reference definition's criteria are conjunctive, final cohort membership is invariant to the order in which they are applied; the order of application affects the intermediate cumulative counts of Table S3 only.

### Fidelity against the values published on the original database

Arm A is a calibration arm, and its value depends on the re-implementations behaving, on this database, like the definitions they re-implement behaved on the original one. Fidelity was therefore assessed by comparing each team's agreement with the reference definition here against the corresponding value published by the reproduction study on its own database and common data model.

**Every published quantity used in this comparison is a value the reproduction study prints, and each is used as printed.** Per-team cohort sizes come from its Supplementary Table 3, row "Total, n". Per-team agreement values come from the data labels of its Figure 4, which prints each team's Jaccard index against the reference implementation to one decimal place; the Results text additionally states two of the nine, Team 8 at 35.4% and Team 5 at 0%. Per-team overlap with the reference cohort is the one quantity the study does not print, but it is fixed by the two it does: since the Jaccard index is the shared count divided by the union, the shared count follows as J × (team size + reference size) ÷ (1 + J). Any such value is reported here as derived rather than published, and inherits only the rounding of the one-decimal agreement percentages — under ±30 patients at the largest cohort size.

### Table S2. Agreement with the reference definition, as published and as obtained here

The two agreement columns are Jaccard indices against the reference definition executed on the same database as the team definitions being scored: the reproduction study's own reference cohort of 6196 patients for the published column, and this study's reference cohort of 17 158 patients for the column obtained here. Rows are ordered by the published value, descending. The last four columns decompose each comparison into the patients a definition shares with its reference cohort and those the reference cohort holds alone; for the published study these are derived rather than printed, as footnote g sets out.

[a] Published as data labels in Figure 4 of the reproduction study, which gives each team's Jaccard index against the reference implementation to one decimal place; the values for Teams 5 and 8 are also stated in that study's Results text. The reproduction study prints these as percentages; they are restated here as fractions on the 0-to-1 scale used throughout this manuscript. Same quantity, different notation.

[b] Rank 1 is the lowest agreement and rank 9 the highest, within each column separately.

[c] Teams 2 and 6 carry the same published value and therefore tie at the mean of ranks 5 and 6.

| Team | Agreement with the reference cohort | | | Rank[b] | | Cohort size | | Shared with the reference cohort | | Reference cohort only | |
|---|---|---|---|---|---|---|---|---|---|---|---|
| | Published[a] | Here | Ratio, here ÷ published | Published | Here | Published[f] | Here | Published[g] | Here | Published[g] | Here |
| Team 8 | 0.354 | 0.508 | 1.44 | 9 | 9 | 12 618 | 31 381 | 4919 | 16 355 | 1277 | 803 |
| Team 9 | 0.172 | 0.300 | 1.74 | 8 | 8 | 31 253 | 45 329 | 5496 | 14 420 | 700 | 2738 |
| Team 3 | 0.161 | 0.259 | 1.61 | 7 | 5 | 11 260 | 14 886 | 2421 | 6589 | 3775 | 10 569 |
| Team 2 | 0.153 | 0.266 | 1.74 | 5.5[c] | 6 | 18 701 | 41 020 | 3304 | 12 220 | 2892 | 4938 |
| Team 6 | 0.153 | 0.285 | 1.86 | 5.5[c] | 7 | 2159 | 9155 | 1109 | 5839 | 5087 | 11 319 |
| Team 1 | 0.128 | 0.210 | 1.64 | 4 | 4 | 43 641 | 73 560 | 5655 | 15 729 | 541 | 1429 |
| Team 7 | 0.094 | 0.189 | 2.01 | 3 | 3 | 63 619 | 83 706 | 5999 | 16 003 | 197 | 1155 |
| Team 4 | 0.008 | 0.013 | 1.59 | 2 | 2 | 8417 | 17 995 | 116 | 441 | 6080 | 16 717 |
| Team 5 | <0.001[d] | 0.000 | —[d] | 1 | 1 | 3126 | 2614 | ~0 | 0 | ~6196 | 17 158 |
| Median | 0.153 | 0.259 | 1.69[e] | | | 12 618 | 31 381 | 3304 | 12 220 | 2892 | 4938 |

[d] Figure 4 prints Team 5's value as less than 0.1% while the Results text gives it as 0%; no ratio is defined either way. Our Team 5 cohort shares no patients at all with the reference cohort.

[e] Median of the eight defined ratios. Ratios are computed from unrounded Jaccard indices, so Team 4's differs slightly from the quotient of the two rounded values printed beside it.

[f] Published in Supplementary Table 3 of the reproduction study (row "Total, n"), and reproduced here as published.

[g] Not printed by the reproduction study. Derived from the two quantities that are, through the Jaccard identity: shared = J × (team n + reference n) ÷ (1 + J), using the reference cohort of 6196 given in that study's main text. Its Supplementary Table 3 gives 6356 instead, which would shift each derived shared count by up to about 42 patients and each derived reference-cohort-only count by 118 to 159. Derived values also inherit the rounding of the published one-decimal agreement percentages, under ±30 patients at the largest cohort size. The corresponding columns for this study are measured directly from the patient sets, not derived.

**Supplementary Figure S1**

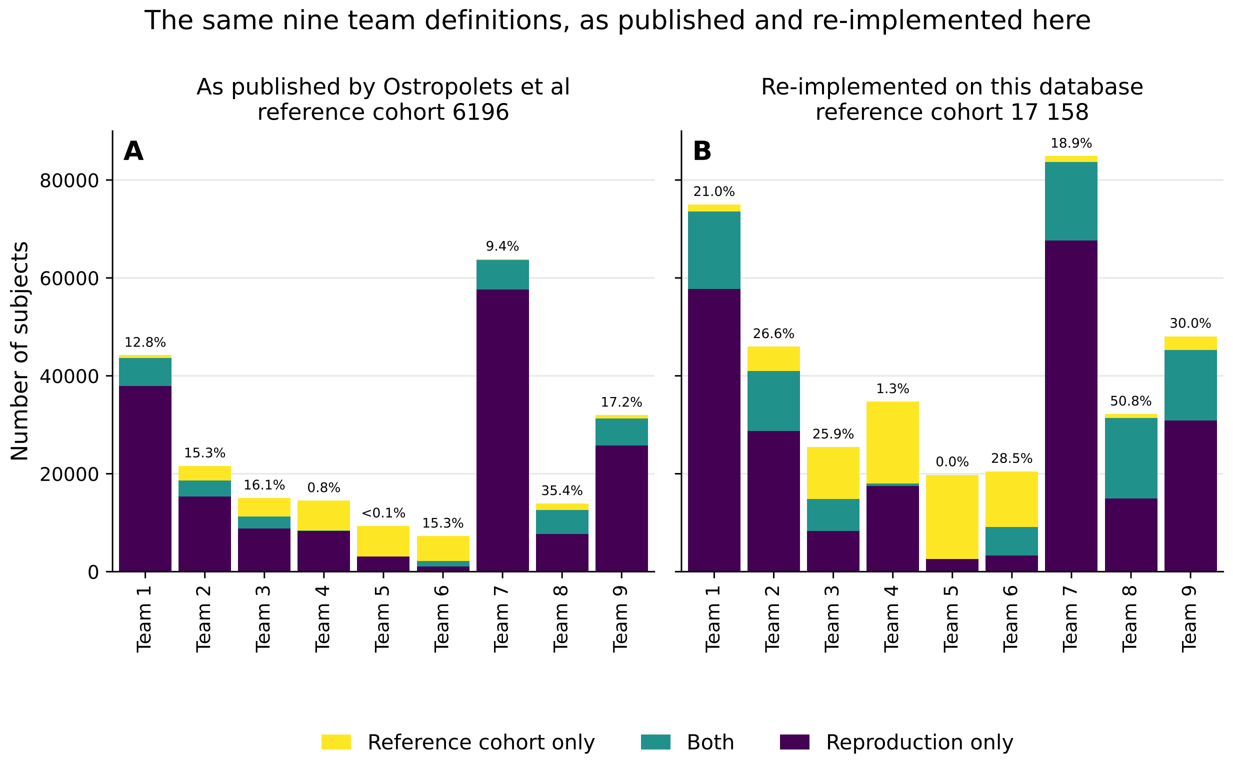


**Figure S1. The same nine team definitions, as published and as re-implemented here.** Each bar is one team's cohort definition and spans every patient in either that team's cohort or the reference cohort it is scored against, so the bar height is the union of the two; the lower two segments sum to the team's own cohort size and the upper two to the reference cohort size. The value above each bar is that definition's Jaccard index against its own panel's reference cohort, expressed as a percentage. (A) The nine team cohorts as published by Ostropolets et al, on that study's database and against its reference cohort of 6196 patients. Cohort sizes are taken from that study's Supplementary Table 3 and agreement values from its Figure 4; the shared segment is not published and is derived from those two quantities through the Jaccard identity, as footnote g to Table S2 sets out. The value for Team 5, published as less than 0.1%, is drawn as zero. (B) The same nine published definitions re-implemented on this study's database and scored against its reference cohort of 17 158 patients; here both the cohort sizes and the shared counts are measured directly from the patient sets. Team numbering is shared between the panels, so Team k is the same team in both. The vertical scale is common to the two panels, but their reference cohorts differ in size, so bar heights are directly comparable only within a panel; the Jaccard index printed above each bar is the quantity intended for comparison across panels.

Alt text: (A) Stacked bar chart, nine team cohorts as published, scored against a reference cohort of 6196 patients. Bars span 7000 to 64 000 patients in either cohort. Shared segments are thin. Agreement printed above each bar runs from below 0.1 percent to 35.4 percent.

Alt text: (B) The same chart for the same nine definitions rebuilt on this study database, scored against a reference cohort of 17 158 patients. Bars span 20 000 to 85 000 patients. Shared segments are visibly thicker. Agreement runs from 0.0 percent to 50.8 percent.

Three conclusions follow.

**Rank order correlates closely, but is not an exact match.** Spearman $\rho$ between the published agreement values and ours is 0.946, and Kendall $\tau$-b is 0.873, with 33 concordant and **2 discordant** of the 35 untied pairs. The two discordant pairs arise from a single reordering, confined to one block of three teams — Teams 2, 3, and 6 — whose published values span 0.8 percentage points (16.1%, 15.3%, and 15.3%) and are therefore not meaningfully ordered in the published data to begin with. Cohort size supplies an independent check on the same question, and one unaffected by the rounding of the published one-decimal agreement percentages: Spearman $\rho$ between the published per-team sizes and ours is 0.967 and Kendall $\tau$-b is 0.889, with 34 of the 36 pairs concordant. The two discordant pairs are Teams 3 and 4, and Teams 5 and 6 — in each case a pair that is adjacent in rank on both databases.

**The uplift is near-constant.** Agreement is higher on this database than on the original for every team with a defined ratio, eight of the nine, by a multiplicative factor between 1.44 and 2.01 with a median of 1.69. The shift is systematic rather than selective: it moves the whole distribution, and it moves it by approximately the same factor at every rank. Its cause cannot be decomposed, because this database differs from the original both in population and in schema and the original database was not accessible to us; that limitation is stated in the manuscript's Discussion. But a uniform multiplicative shift is what a database effect looks like, and is not what a translation error looks like: a translation error would displace the definitions it affected and leave the others in place. Cohort sizes rise as well, by a median factor of 1.69, but less uniformly than agreement (range 0.84 to 4.24): Team 5 is slightly smaller on this database and Team 6 roughly four times larger. The calibration rests on agreement, which is the more stable of the two quantities.

**Both extremes are preserved, by the same two teams.** The published range runs from below 0.1% (Team 5) to 35.4% (Team 8). Our range runs from 0.000 (Team 5) to 0.508 (Team 8). The same two teams anchor both ends of the distribution on a different database, under an independent re-implementation built from the published specifications alone.

One further consequence of reading these values is worth recording, because it bears on a benchmark named in the manuscript. The nine published per-team values have a median of 0.153, not the 0.094 the reproduction study states in its abstract and results. The value 0.094 is exactly that study's Team 7 value, and the interquartile range published alongside it — 15.3% to 16.2% — does not contain 9.4%, so the published sentence cannot be internally consistent. The manuscript reports the same-database Arm A median of 0.259

as its primary benchmark and names both published figures as context.

#### Two definitions whose insulin criteria are named as exclusions but coded as requirements

Two of the nine team definitions contain a criterion named as an insulin exclusion whose compiled logic requires insulin. Both were reproduced as coded.

In Team 4, the criterion “No insulin prior to index date” compiles to a group requirement of at least one insulin exposure at any time up to and including the index date — HAVING COUNT >= 1, against the = 0 used by every genuine exclusion in the same definition. In Team 5, the criterion “Insulin or type 1 diabetes” is expressed as “at most 0” over three individually absent criteria — insulin as a drug, insulin as a device, and type 1 diabetes. That compiles to “at most zero of these are absent”, which is to say all three present. The definition therefore requires insulin and type 1 diabetes rather than excluding them. On this database, insulin as a drug and insulin as a device resolve to the same dispensing record, so the two insulin arms collapse into one.

The reproduction study’s own supplementary deviation table labels both criteria as insulin *exclusions*, in the same direction as the reference definition. That label cannot be right, and it is the published results themselves that show it:

1. The reference definition excludes insulin in the 365 days before the index date.

2. If Team 5 also excluded insulin, its cohort would necessarily overlap the reference cohort substantially.

3. The reproduction study reports Team 5’s agreement with the reference as 0% in its text and as below 0.1% on its figure, while its Supplementary Table 3 reports a Team 5 cohort of 3126 patients. The cohort is therefore not near-empty; it is somewhere else entirely.

4. A cohort that is both non-empty and essentially disjoint from a no-insulin cohort is an insulin-requiring cohort. No other reading produces it.

5. Team 4 falls in the same regime for the same reason: given the reference definition’s insulin exclusion, its published agreement of 0.8% is only producible under a require-insulin reading.

The re-implementations reproduce both results. Team 5’s cohort here shares zero patients with the reference cohort and zero patients with each of the other eight team cohorts — completely disjoint — against a published value of 0% in the text and below 0.1% on the figure. Team 4’s cohort intersects the reference cohort in 441 patients, a Jaccard index of 0.013, against a published 0.8%. The reproduction study’s report of one team reaching zero agreement is recoverable only under the as-coded reading. Under the as-named reading, that team’s cohort would overlap the reference cohort substantially and the published result could not be reproduced at all.

Two further criteria in Team 5’s definition are misnamed in the same way and were likewise reproduced as coded: its “Monotherapy” criterion in fact requires at least one antidiabetes drug in the 365 days before the index date, and its “Pregnancy” criterion evaluates the index date alone rather than a window around it.

#### Other fidelity controls

The concept sets published with the ten definitions are semantically identical across all ten, verified file by file, so cohort logic is the only variable in this arm. The reference definition’s attrition ladder was compared criterion by criterion against the percentages published for the corresponding criteria on the original database; that comparison is Table S3.

### SUPPLEMENTARY METHODS 2 — AGENT INSTRUCTION (ARM B)

The text reproduced below is the complete instruction issued to the nine Arm B agent runs. It was delivered as the single substantive instruction to each of those runs, and it was byte-identical across all nine Arm B runs. Nothing else was said to any Arm B run.

The instruction is reproduced here verbatim and in full; line wrapping reflects the width of this page.

> Using the concept sets already provided in your codebook and the analytic tools available to you, reproduce the target cohort — the new-user GLP-1 receptor agonist arm — defined in the excerpts below from the Albogami et al. paper and figures 1 & 2 from the supplementary materials (/workspace/albogami_figures/), on this dataset. Every codeset in your codebook is a frozen concept set (in /workspace/.codebook/) from Albogami's study: use them as given, do not re-derive or modify them — only the cohort logic is yours to determine. Visit types (inpatient, emergency, outpatient) are not codesets — they are represented by the dataset's visit fields (e.g., ‘location_of_care‘, ‘inpatient_ind‘) described in the schema. You must try to reproduce the GLP-1RA cohort on this dataset — exactly as described — by defining the inclusion and exclusion criteria, and order of application, based on the description. You must present a protocol for my approval. Produce the final set of ‘patient_id‘s in the target cohort, plus a brief summary of the criteria you applied and in what order.
>
> Albogami Extracts:
>
> Research design and methods
>
> Study design and data source
>
> We assembled a cohort of new users of glucagon-like peptide 1 receptor agonists (GLP-1RA) among adults with comorbid type 2 diabetes (T2D) and chronic lower respiratory disease (CLRD), using a new-user, active-comparator design with dipeptidyl peptidase-4 inhibitors (DPP-4I) as the comparator class. Analyses used longitudinal administrative claims data with both medical and pharmacy benefits, comprising diagnoses and procedures from inpatient and outpatient encounters and outpatient pharmacy dispensings. Diagnoses were ascertained from diagnosis codes and medications from their dispensing records, using predefined concept sets for each clinical entity. The study timeline and the windows over which each criterion was assessed are shown in Figure 1, and the sequence of cohort-selection criteria is shown in Figure 2. All windows described below are defined relative to the index date, and every criterion was evaluated within the study period shown in Figure 1.
>
> Cohort entry and exposure
>
> Cohort entry was defined by initiation of a GLP-1RA. Because fixed-dose combinations of a GLP-1RA with insulin represent insulin exposure, only GLP-1RA products that did not contain insulin were treated as qualifying exposures. The index date was the date of the earliest such GLP-1RA dispensing that was itself preceded by at least 365 days of continuous health-plan enrollment, provided as a single, unbroken enrollment period in both the medical and the pharmacy benefit; a dispensing not meeting this baseline requirement did not serve as the index event, and a patient entered the cohort on the earliest dispensing that did. To reinforce the new-user design, we further excluded any patient with a GLP-1RA dispensing in the 365 days before the index date. Patients were required to be older than 17 years of age at the index date.
>
> Comorbid disease requirements
>
> Eligible patients had evidence of both T2D and CLRD during the baseline period. T2D was established by at least one type 2 diabetes diagnosis in the 365 days up to and including the index date. CLRD was established, within the same 365 days up to and including the index date, by any one of three criteria: at least one CLRD diagnosis recorded during an inpatient encounter; CLRD diagnoses on at least two distinct calendar dates; or at least one CLRD diagnosis together with at least one dispensing of a medication used to treat CLRD. A CLRD medication in the absence of a

CLRD diagnosis did not, by itself, establish CLRD.

Add-on therapy requirement

Because GLP-1RA were studied as add-on therapy to an existing antidiabetes regimen, each patient was required to be receiving concurrent antidiabetes therapy at the index date. This was operationalized as a dispensing of an antidiabetes drug before the index date whose days' supply extended to at least the index date, so that the patient was actively treated with another antidiabetes medication when GLP-1RA therapy began.

Exclusion criteria

Patients were excluded if, in the 365 days before the index date, they had a diagnosis of type 1 diabetes; of cystic fibrosis, lung cancer, pulmonary embolism, or pulmonary hypertension; of thyroid carcinoma; or of any condition that frequently requires chronic systemic corticosteroid therapy, other than chronic obstructive pulmonary disease and asthma. Women were excluded for pregnancy if they had a pregnancy-related diagnosis within 180 days before or after the index date. To preserve the new-user, active-comparator design, patients were excluded if they had any DPP-4I dispensing in the 365 days up to and including the index date, or any insulin dispensing in the 365 days before the index date.

Cohort

Patients who satisfied every inclusion requirement and none of the exclusion criteria constituted the GLP-1RA new-user cohort (Figure 2).

Two deliberate departures from the human condition are recorded in the manuscript and repeated here. First, the instruction directs the agent to reproduce the cohort "exactly as described" and to derive the criteria and their order from the description; this is marginally more fidelity-directive than the wording given to the human teams, who were asked to define any number of criteria in any order. Second, no time limit was imposed, whereas the human teams were given a time limit of eight hours. Nonetheless, the agents completed cohort construction in less than one hour.

### SUPPLEMENTARY METHODS 3 — INPUT BUNDLE AND ACCOUNT ISOLATION

#### Placed in each account

Each of the nine Arm B accounts was provisioned identically, before any run, with the following.

**The target study in full.** The complete published paper and its complete supplement, as two PDF files in the agent's working directory at /workspace/albogami/ (Albogami.pdf and Albogami_supplement.pdf). The files were byte-identical in all nine accounts. The materials were supplied whole, not as curated extracts, so that each agent had access to the same free-text description the human teams worked from.

**The frozen code set library.** Twenty code set files installed in the agent's code library at /workspace/.codebook/. Every file is a concept set from the same library the human teams were given. Codes were fixed and the instruction forbids re-deriving or modifying them, so cohort logic was the only variable.

**The database and its documentation.** The HealthVerity Base claims database with its schema and data dictionary, together with the agent's own analytic tools. This is the analogue of the human teams' native competence in the common data model and cohort-building environment they used.

#### Withheld from every account

Nothing was supplied beyond the two PDFs and the code set library. Specifically withheld were the reproduction study that motivated this work, the reference cohort's size and hash, the definitions produced by any human team, all intermediate results of this study, and any information from any other run. Disclosing the target would have voided the run.

The reference cohort size and hash appear nowhere in the target study's materials, so the target was never disclosed even implicitly. The target study does report its own cohort counts in a supplementary figure, but those counts were computed on the target study's own data rather than on the database used here, and the human teams saw the same figure.

#### The frozen code set library

The library comprises 20 code set files, each one a concept set supplied with the target study's published definitions. Two of them, a CLRD drug list and a corticosteroid-indication list, have no published basis in the target study itself, but both were available to the human teams through the original cohort-building environment and so belong in a faithful reconstruction. A twenty-first file present in the general provisioning bundle — a partition of the antidiabetes drug code set by drug ingredient — was to be removed from the Arm B and Arm C accounts by an explicit setup precondition. That file is an artefact of this study's own deterministic Arm A re-implementations rather than a concept set of the target study; leaving it in place would have contradicted the instruction's statement that every code set in the codebook comes from the target study's cohort-building environment, and could have prompted an agent to decompose antidiabetes drugs by ingredient in a way the target study never did. The setup procedure therefore specified a codebook holding the 20 target-study code set files only.

The partition exists because OMOP drug eras are defined per RxNorm ingredient while the supplied code sets are at National Drug Code level, so two of the published team rules cannot be expressed without an ingredient grain: Team 4 requires at least two distinct antidiabetic ingredients whose eras span the index date, and Team 2 requires at least one ongoing ingredient era. Three Arm A re-implementations load it — those of Teams 2, 4 and 9, the last to resolve dipeptidyl peptidase-4 inhibitors, concept set 8 being a strict subset of the antidiabetes set. The reference definition does not use it, and neither do the six remaining team definitions, so withholding it from the Arm B and Arm C accounts left nothing about the reference cohort unreproducible.

#### Omission of the three visit-type concept sets

The codebook is the target study's concept-set pool minus its three visit-type concept sets — inpatient or inpatient-with-emergency, emergency, and outpatient. Those three were omitted because the database used here represents care setting as fields on its visit records rather than as code lists, so a visit-type code set has no equivalent in this schema. To prevent an agent searching for a concept set that does not exist, the instruction states explicitly that visit types are not code sets and points the agent to the relevant visit fields by name. The three omitted concept sets are thus supplied through the schema rather than through the codebook, and no visit-type information was withheld.

#### Account isolation

Each of the nine Arm B runs was executed in a separate account, giving each an independent sandboxed workspace and an independent memory. A run had no memory of any other run and no access to any other run's information, and no cross-run information was introduced at any point. The nine accounts are the analogue of the nine human teams. Arm C was provisioned on the same model, and its nine runs were executed across nine accounts.

#### Deliverables captured from each run

Two artefacts were captured from every run for scoring: the final set of patient identifiers in the cohort, scored against the reference cohort by the Jaccard index; and the run's own brief summary of the inclusion and exclusion criteria it applied and the order in which it applied them, used to characterise where each run departed from the reference definition.

### SUPPLEMENTARY METHODS 4 — REVISED SPECIFICATION (ARM C)

Arm C replaced the target study's published paper and supplement with a rewritten methods section, reproduced in full below. The rewrite restates the same cohort definition as continuous prose in the original's voice and structure, with every internal contradiction in the published account resolved to a single reading. Where the published paper's text, tables, and figures disagreed — most consequentially on how chronic lower respiratory disease is ascertained and on the length of the prior-exposure washout — the rewrite states one rule and states it once.

The rewritten methods section names no database, discloses no target cohort count, and makes no reference to this study. It was delivered inside the same instruction wrapper used for Arm B, which names Albogami et al. as the source of the excerpts it introduces. Arm C agents therefore knew which study they were reproducing, exactly as Arm B agents did; what differed was the material supplied with that wrapper, the rewritten methods and two count-free companion figures in place of the paper and its supplement. Because it describes the reference cohort as it was actually built, Arm C measures convergence on a described target rather than recovery of an undisclosed one; the manuscript's Discussion sets out what this does and does not license.

The rewritten methods section was supplied together with two count-free companion figures, redrawn to match it: a study timeline showing the windows over which each criterion is assessed, and a cohort-selection flow chart showing the sequence of criteria. Both are referenced in the text below as Figure 1 and Figure 2 respectively, and are reproduced here as Figures S2 and S3. Neither carries any patient count, so neither discloses the target. The rewritten methods and its two figures replaced the two source PDFs supplied to Arm B, whose nine runs were executed across nine accounts. The frozen code set library was the same library of 20 files supplied to Arm B.

#### Revised specification, reproduced in full as supplied

The text reproduced below is the complete instruction issued to the nine Arm C agent runs. It corrects the ambiguities and contradictions in the target study to align with the computer code used to construct the reference cohort, was delivered as the single substantive instruction to each of those runs, and it was byte-identical across all nine Arm C runs. Nothing else was said to any Arm C run.

The instruction is reproduced here verbatim and in full; line wrapping reflects the width of this page.

> Using the concept sets already provided in your codebook and the analytic tools available to you, reproduce the target cohort — the new-user GLP-1 receptor agonist arm — defined in the excerpts below from the Albogami et al. paper and figures 1 & 2 from the supplementary materials (/workspace/albogami_figures/), on this dataset. Every codeset in your codebook is a frozen concept set (in /workspace/.codebook/) from Albogami's study: use them as given, do not re-derive or modify them — only the cohort logic is yours to determine. Visit types (inpatient, emergency, outpatient) are not codesets — they are represented by the dataset's visit fields (e.g., ‘location_of_care‘, ‘inpatient_ind‘) described in the schema. You must try to reproduce the GLP-1RA cohort on this dataset — exactly as described — by defining the inclusion and exclusion criteria, and order of application, based on the description. You must present a protocol for my approval. Produce the final set of ‘patient_id‘s in the target cohort, plus a brief summary of the criteria you applied and in what order.
>
> Albogami Extracts:
>
> Study design and data source
>
> We assembled a cohort of new users of glucagon-like peptide 1 receptor agonists (GLP-1RA) among adults with comorbid type 2 diabetes (T2D) and chronic lower respiratory disease (CLRD), using a new-user, active-comparator design with dipeptidyl peptidase-4 inhibitors (DPP-4I) as the comparator class. Analyses used longitudinal administrative claims data with both medical and pharmacy benefits, comprising diagnoses and procedures from inpatient and outpatient encounters and outpatient pharmacy dispensings. Diagnoses were ascertained from diagnosis codes and medications from their dispensing records, using predefined concept sets for each clinical entity. The study timeline and the windows over which each criterion was assessed are shown in Figure 1, and the sequence of cohort-selection criteria is shown in Figure 2. All windows described below are defined relative to the index date, and every criterion was evaluated within the study period shown in Figure 1.
>
> Ascertainment conventions. Diagnoses were matched only on records qualified as ICD-10 (i.e., whose diagnosis-vocabulary qualifier is ICD-10); each code was matched verbatim against the predefined concept set as provided, with no reformatting of punctuation. No cohort criterion uses procedure codes — every match is against a diagnosis record or a dispensing (pharmacy) record. Each clinical event was counted only if its event date (a diagnosis's start date, a dispensing's fill date) fell within the study data window of 1 January 2020 through 31 October 2025; the baseline-enrollment check that defines index eligibility (below) is exempt from this lower bound. Enrollment records were classified by benefit type by substring on the lower-cased benefit-type value: a record counted as a medical benefit if that value contained 'medical', and as a pharmacy benefit if it contained 'pharm' or 'rx'; a single enrollment record may satisfy both benefits. A visit was treated as an inpatient encounter if and only if its inpatient indicator was truthy — the indicator value, lower-cased, being one of y, yes, true, t, or 1 — with no additional place-of-service or care-location requirement. A patient was treated as female only if the recorded sex was female; a missing or unknown sex was treated as not female. Age at the index date was computed as the index year minus the year of birth; a patient with a missing or non-numeric year of birth failed the age criterion. All inclusion and exclusion criteria were combined with AND, and the order in which they are applied does not affect cohort membership.
>
> Cohort entry and exposure
>
> Cohort entry was defined by initiation of a GLP-1RA. Because fixed-dose combinations of a GLP-1RA with insulin represent insulin exposure, only GLP-1RA products that did not contain insulin were treated as qualifying exposures. Candidate index dispensings were the insulin-free GLP-1RA dispensings occurring between 1 January 2021 and 31 October 2025 (this eligibility window begins one year inside the data window, so that a full baseline year is available for every candidate; it is the binding lower bound for entry). A candidate dispensing qualified only if it was itself preceded by at least 365 days of continuous health-plan enrollment in both the medical and the pharmacy benefit, where the 365-day requirement in each benefit was met by a single enrollment record — one uninterrupted record whose coverage began at least 365 days before the dispensing and extended through the dispensing date. Adjacent or overlapping enrollment records were not merged, stitched, or gap-consolidated into a longer period; the requirement had to be satisfied by one record per benefit. The index date was the earliest qualifying dispensing: if a patient's earliest insulin-free GLP-1RA dispensing did not qualify, the earliest later dispensing that did was used instead, and a patient with no qualifying dispensing did not enter the cohort. To reinforce the new-user design, we further

excluded any patient with a dispensing of the same insulin-free GLP-1RA products in the 365 days before the index date. Patients were required to be older than 17 years of age at the index date.

Comorbid disease requirements

Eligible patients had evidence of both T2D and CLRD during the baseline period. T2D was established by the presence of at least one type 2 diabetes diagnosis in the 365 days up to and including the index date (a single qualifying diagnosis suffices; there is no inpatient/outpatient encounter-count requirement). CLRD was established, within the same 365 days up to and including the index date, by any one of three criteria: at least one CLRD diagnosis recorded on an inpatient encounter (a visit whose inpatient indicator is true — an outpatient CLRD diagnosis together with a separate inpatient visit does not satisfy this arm); CLRD diagnoses on at least two distinct calendar dates in any setting; or at least one CLRD diagnosis together with at least one dispensing of a medication used to treat CLRD. A CLRD medication in the absence of a CLRD diagnosis did not, by itself, establish CLRD.

Add-on therapy requirement

Because GLP-1RA were studied as add-on therapy to an existing antidiabetes regimen, each patient was required to be receiving concurrent antidiabetes therapy at the index date. This was operationalized as a dispensing of any drug in the full antidiabetes-drug concept set (used as provided, without pruning) whose dispensing date fell before the index date and whose days' supply reached the index date, so that the patient was actively treated with another antidiabetes medication when GLP-1RA therapy began. A dispensing on day f with s days' supply was taken to cover the interval from f through $f + s - 1$; the drug was active at index if f fell before the index date and $f + s - 1$ fell on or after it. A missing or non-numeric days' supply was treated as zero.

Exclusion criteria

Patients were excluded if, in the 365 days before the index date, they had a diagnosis of type 1 diabetes; of cystic fibrosis, lung cancer, pulmonary embolism, or pulmonary hypertension; of thyroid carcinoma; or of any condition that frequently requires chronic systemic corticosteroid therapy, other than chronic obstructive pulmonary disease and asthma (this carve-out is already reflected in the frozen concept set, which is applied verbatim — no codes are added or removed). Women were excluded for pregnancy if they had a pregnancy-related diagnosis within 180 days before or after the index date; this criterion used the pregnancy diagnosis-code concept set only — pregnancy procedure and observation codes were not used — and applied to female patients only. To preserve the new-user, active-comparator design, patients were excluded if they had any DPP-4I dispensing in the 365 days up to and including the index date, or any insulin dispensing in the 365 days before the index date.

Cohort

Patients who satisfied every inclusion requirement and none of the exclusion criteria constituted the GLP-1RA new-user cohort (Figure 2).

### Supplementary Figure S2

Figure S2. Study design and timeline of baseline assessment windows, as supplied to Arm C. The first of the two count-free companion figures issued with the revised specification, and numbered Figure 1 within that package. Each bar is the window, in days relative to the index date, over which one criterion is assessed; the vertical line marks the index date.

Alt text: Timeline chart. Eight criteria appear as horizontal bars measured in days relative to the index date, which is marked by a vertical line at day 0. Continuous enrollment, type 2 diabetes, chronic lower respiratory disease and the comparator washout span the 365 days up to and including index. The exposure washout and the medical condition exclusions span the 365 days before index. Pregnancy exclusion spans 180 days either side of index. Add-on antidiabetes therapy is an arrow reaching day 0. A key distinguishes inclusion, exclusion, entry and add-on criteria.

Figure 1. Study design and timeline of baseline assessment windows

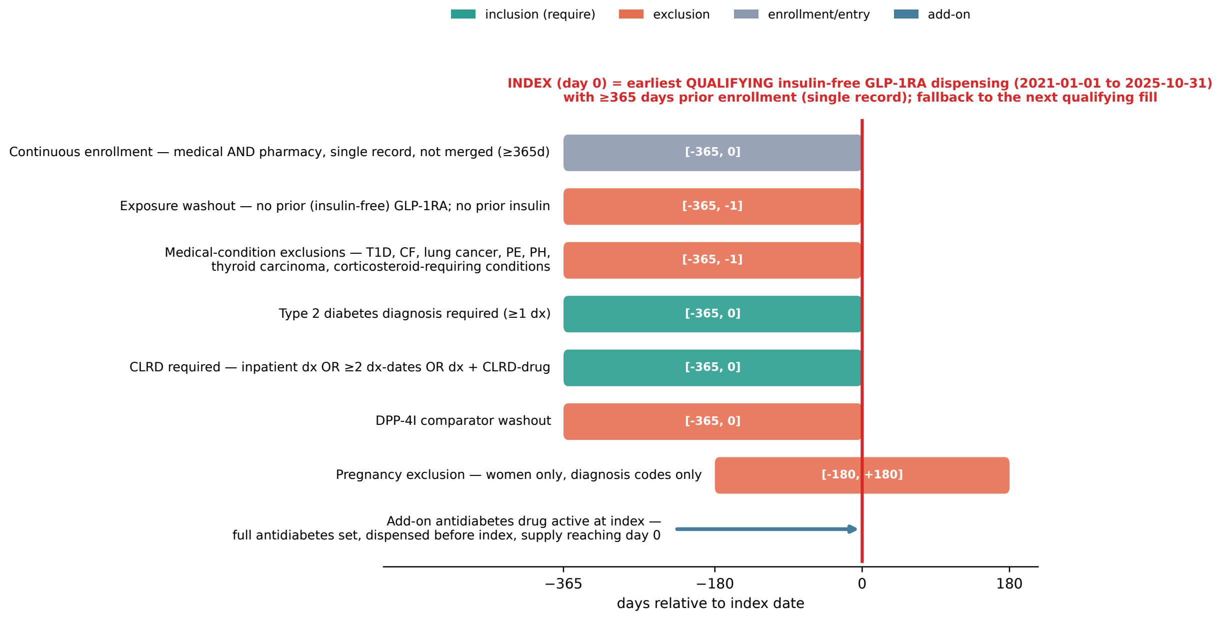


*Study period: index dates 2021-01-01 to 2025-10-31; clinical events (diagnoses, dispensings) assessed within 2020-01-01 to 2025-10-31. Day 0 = index. Exclusion windows end the day before index; the type 2 diabetes, CLRD, and DPP-4I windows include the index date. Diagnoses use ICD-10-qualified records, matched verbatim; pregnancy uses diagnosis codes only. All criteria are applied jointly (AND). Enrollment benefit type by substring: medical = value contains 'medical'; pharmacy = contains 'pharm' or 'rx' (one record may satisfy both).*

#### Supplementary Figure S3

Figure S3. Cohort selection, as supplied to Arm C. The second companion figure, numbered Figure 2 within that package: cohort entry, the inclusion requirements that must all hold, the exclusions of which none may hold, and the resulting new-user cohort.

Alt text: Flow diagram. Cohort entry is the earliest qualifying insulin-free glucagon-like peptide-1 receptor agonist dispensing with at least 365 days of continuous medical and pharmacy enrollment. Two boxes below it list the criteria applied. Include only if all hold covers adult age, type 2 diabetes, chronic lower respiratory disease and add-on antidiabetes therapy. Exclude if any hold covers prior exposure, type 1 diabetes, cystic fibrosis, lung cancer, pulmonary embolism, pulmonary hypertension, thyroid carcinoma, conditions requiring chronic corticosteroids, pregnancy, and prior comparator or insulin use. Both lead to the final new-user cohort.

### SUPPLEMENTARY TABLE S3 — REFERENCE COHORT ATTRITION

#### Table S3. Attrition of the reference cohort, by criterion, with corresponding published percentages

The denominator for every percentage in the "Satisfying" column is the index population — the first in-window GLP-1RA dispensing with at least 365 days of prior medical and pharmacy coverage — which is 747 046 patients, taken as 100%. "Satisfying" is the marginal count: the number of index patients meeting that one criterion considered alone. "Cumulative n" is the running intersection, that is, the attrition ladder. Rows follow the order in which the rules are applied; the "Paper criterion" column gives the corresponding criterion number in the published study, which numbers them differently. Because all twelve of the reference

Figure 2. Cohort selection

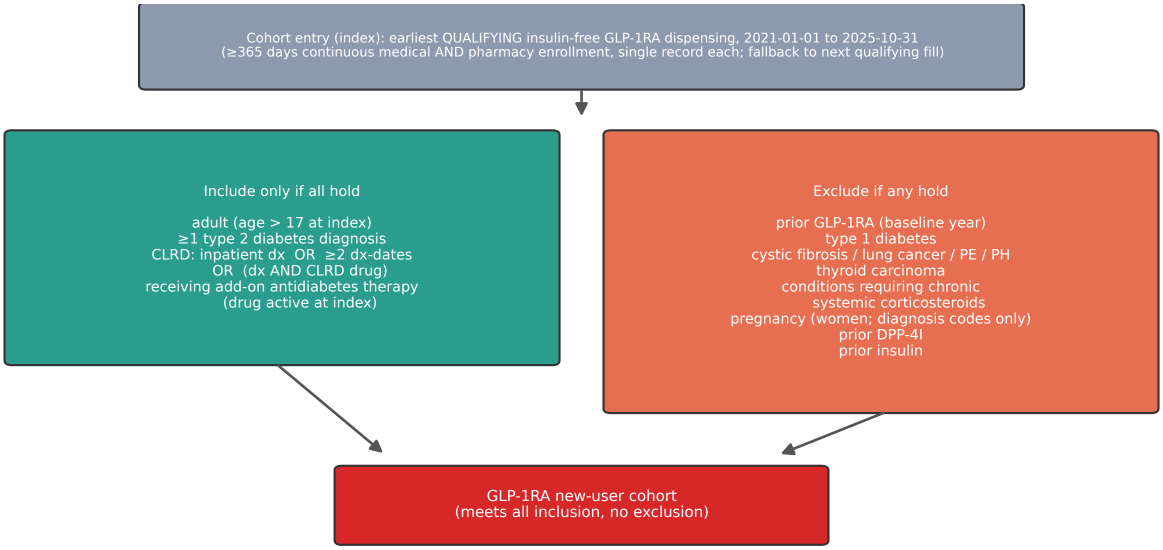


*All criteria are applied jointly (AND); order of application does not affect cohort membership.*
*Each concept is ascertained from its predefined set; diagnoses use ICD-10-qualified records; pregnancy uses diagnosis codes only.*

| Paper criterion | Criterion as implemented | Satisfying, n (%) | Cumulative n | Published, %[a] |
|---|---|---|---|---|
| Entry | idx — first in-window GLP-1RA dispensing (cs4) with at least 365 days of prior medical and pharmacy coverage | 747 046 (100.0) | 747 046 | 100 (entry) |
| 1 | r0 — no prior GLP-1RA (cs4) in the 365 days before index (new user) | 522 301 (69.9) | 522 301 | 98.7[b] |
| 3 | r1 — age older than 17 years | 738 812 (98.9) | 515 116 | 99.9 |
| 6a | r2 — no type 1 diabetes (cs16) in the 365 days before index | 721 472 (96.6) | 500 203 | —[c] |
| 4 | r3 — type 2 diabetes (cs18) in the 365 days up to and including index | 542 811 (72.7) | 330 157 | 75.4 |
| 5 | r4 — CLRD (cs19) in the 365 days up to and including index: inpatient diagnosis, or diagnoses on at least two distinct days, or a diagnosis with a CLRD drug (cs6) | 149 234 (20.0) | 70 323 | 7.8[d] |
| 6b | r5 — no cystic fibrosis, lung cancer, pulmonary embolism, or pulmonary hypertension (cs48, cs43, cs39, cs40) in the 365 days before index | 726 771 (97.3) | 64 756 | —[c] |
| 6c | r6 — no thyroid carcinoma (cs42) in the 365 days before index | 744 092 (99.6) | 64 441 | —[c] |
| 6d | r7 — no corticosteroid-requiring conditions (cs45) in the 365 days before index | 589 830 (79.0) | 44 151 | —[c] |
| 7 | r8 — not pregnant (cs50) within 180 days either side of index, females | 732 194 (98.0) | 43 249 | 99.2 |
| 9 | r9 — no DPP-4 inhibitor (cs8) in the 365 days up to and including index | 683 310 (91.5) | 38 303 | 83.2 |
| 8 | r10 — no insulin (cs37) in the 365 days before index | 559 345 (74.9) | 29 560 | 70.5 |
| 10 | r11 — active add-on antidiabetes drug (cs10) at index | 423 245 (56.7) | 17 158 | 56.2 |
| — | All criteria: final reference cohort | 17 158 (2.3) | 17 158 | 1.1 |

definition's criteria are conjunctive, final cohort membership is invariant to the order of application; the order affects the intermediate cumulative counts only. Code set identifiers (cs4, cs6, and so on) are those of the frozen code set library.

[a] Percentages published for the corresponding criterion in the reproduction study, computed on a different claims database and common data model. **The two percentage columns are not strictly comparable, because they use different denominators.** Ours is the index population, which already requires 365 days of prior medical and pharmacy coverage; the published percentages are computed before that coverage requirement is applied, which the published study reports as a separate criterion 2 satisfied by 55.3% of its own denominator. Coverage is folded into the index definition here and so has no row of its own.

[b] The marginal values differ because the two studies index differently, not because the new-user rule differs. The published study indexed on first-ever exposure across the much longer window reported in its Table 1, so almost every index event is trivially a new-user event. We index on the first dispensing within a shorter window and enforce the new-user requirement through the 365-day washout, so in-window first dispensings preceded by an earlier one are dropped at this row. The final new-user definition — no prior exposure in the preceding 365 days, with at least 365 days of coverage — is equivalent in the two studies.

[c] The published study reports its criterion 6 as a single grouped row, satisfied by 85.7%, covering type 1 diabetes, cystic fibrosis, lung cancer, pulmonary embolism, pulmonary hypertension, thyroid carcinoma, and corticosteroid-requiring conditions together. Our implementation splits it into the four explicit rules shown here as 6a to 6d, which are therefore not individually comparable with the single published value. A secondary analysis collapsing the four rules gives 562 864 of 747 046, or 75.3%, passing all four jointly, against the published 85.7%: criterion 6 is more restrictive on this database, driven by a higher prevalence of corticosteroid-requiring conditions.

[d] CLRD is markedly more prevalent in this database's index population than in the published study's, at 20.0% against 7.8%. This is the single largest per-criterion divergence between the two, and is consistent with the published study identifying CLRD ascertainment as a leading driver of between-team deviation.

Abbreviations: CLRD, chronic lower respiratory disease; DPP-4, dipeptidyl peptidase-4; GLP-1RA, glucagon-like peptide-1 receptor agonist.

### SUPPLEMENTARY TABLE S4 — RUN TIME PER REPRODUCTION

#### Table S4. Run time (minutes) per reproduction, by arm

| Reproduction | Arm A | Arm B | Arm C |
|---|---|---|---|
| 1 | 17.1 | 29.6 | 49.9 |
| 2 | 17.2 | 30.4 | 53.1 |
| 3 | 14.0 | 25.2 | 38.7 |
| 4 | 19.1 | 28.0 | 50.0 |
| 5 | 13.3 | 36.2 | 42.8 |
| 6 | 16.0 | 21.6 | 48.8 |
| 7 | 18.0 | 24.4 | 35.1 |
| 8 | 16.1 | 17.1 | 36.8 |
| 9 | 11.1 | 31.9 | 36.1 |
| Mean | 15.8 | 27.2 | 43.5 |
| Median | 16.1 | 28.0 | 42.8 |
| 95% CI | 13.8–17.7 | 22.7–31.6 | 38.1–48.9 |

Wall-clock time from the start of each reproduction to the emission of its final cohort, in minutes. Reproduction numbers index independent runs within an arm and are not comparable across arms. Arm A times are for the platform executing the re-implemented human team definitions and are not the eight hours the original teams were given; they are reported for completeness, not as a human-versus-agent comparison. Arm C times are for the nine runs given the de-contradicted specification. Confidence intervals are t-distribution intervals on the arm mean (8 degrees of freedom).
